# Identification of Persistent Radiomics Feature Co-occurrence Across Diverse Tissue Types and Individuals: A Network-Based Analysis of the RADAPT CT Atlas

**DOI:** 10.64898/2026.07.17.26358252

**Authors:** Sajad Amiri, Pardis Afshar, Mohammad Hossein Rohban

## Abstract

**Objectives:** Radiomics pipelines extract hundreds of quantitative features that are widely known to be redundant, but the structure of this redundancy is usually treated as a per-dataset nuisance to be pruned away. We tested the alternative hypothesis that a substantial number of feature–feature correlations are universal: they persist across patients and across anatomically distinct structures because they reflect shared mathematical and image-statistical properties of how the image is summarised, rather than properties of the tissue being imaged.

**Materials and Methods:** We re-analysed the publicly available Radiomics Atlas Dataset of normal Abdominal and Pelvic CT (RADAPT), restricting the analysis to the 526 non-contrast-enhanced examinations of the 531-subject atlas and to the 107 original (non-filtered) PyRadiomics features. The 53 segmented structures were grouped into four broad anatomical categories — bones, muscles, vessels, and parenchymal organs. RADAPT is distributed as one Excel file per structure, with patients as rows and features as columns. Within each structure file we z-score-normalised every feature across patients, computed the absolute Spearman correlation matrix, and retained edges with |ρ| ≥ τ for τ ∈ {0.70, 0.80, 0.90}. We then intersected the edge sets across all structure files to obtain a “universal” correlation graph, in which an edge survives only if it exceeds the threshold in every structure (each estimated across the full patient sample). Stable feature communities were defined as the maximal cliques of this graph. Robustness to patient sampling was tested by repeating the entire pipeline on five independent random splits of each file into two patient halves (10 sub-cohorts per threshold), and the implementation was independently reproduced in R.

**Results:** Despite the strictness of the global-intersection criterion, 34, 24, and 14 stable feature communities survived at τ = 0.70, 0.80, and 0.90 respectively, with the largest cliques containing six members at τ = 0.70 and τ = 0.80 and five members at τ = 0.90. The community structure was clearly interpretable: separate cliques captured (i) variance-like intensity dispersion, (ii) long-run / low-frequency (coarse) texture, (iii) high gray-level texture, (iv) low gray-level texture, (v) volume and surface shape, and (vi) local-homogeneity and energy/entropy duals. On random-half resampling the exact-match recovery rate of these communities was 81.5 %, 86.7 %, and 80.7 % across the three thresholds; departures from exact recovery were almost always a single boundary feature added or dropped, consistent with finite-sample fluctuation of near-threshold edges rather than structural instability. The R re-implementation reproduced the Python results exactly.

**Conclusion:** A substantial portion of radiomics feature collinearity is universal across patients and tissues. We distinguish two layers within it: trivial near-algebraic duals that are universal by construction, and non-trivial cross-matrix-family communities that are the genuine empirical finding. Together they provide an interpretable, definition-grounded basis for aggressive dimensionality reduction, for retrospectively reconciling apparently different feature selections in the literature, and for moving radiomics pipelines toward organ-agnostic, more reproducible models.

**Clinical relevance statement:** Selecting a single representative feature from each universal community shrinks the original-feature space by roughly an order of magnitude without sacrificing biologically distinct information. For example, the five variance-family members (first-order Variance, GLCM SumSquares, GLCM ClusterTendency, GLDM and GLRLM GrayLevelVariance) can be replaced by a single representative, removing redundant degrees of freedom that would otherwise inflate model variance; and labelling each retained feature by its community lets two studies that selected different variance-family names be recognised as having found the same signal, simplifying model development and improving cross-cohort generalisability in clinical CT workflows.

**Key points:**

- Most studies treat radiomics feature collinearity as a per-dataset problem; we show that a substantial portion of it is universal across anatomy.
- Using a global-intersection network on the RADAPT atlas (526 non-contrast examinations), 34, 24, and 14 stable feature communities survive at Spearman thresholds of 0.70, 0.80, and 0.90.
- Random-half resampling and a cross-language (Python and R) re-implementation confirm that these communities are reproducible properties of the feature extractor, not of any single patient subset.

## 1. Introduction

Radiomics converts standard radiological images into hundreds of quantitative descriptors of intensity, shape, and texture, and has rapidly become a cornerstone of imaging-biomarker research and image-based machine learning [1, 2, 3]. Public standardisation efforts such as the Image Biomarker Standardisation Initiative (IBSI) and the open-source PyRadiomics package have made these descriptors reproducible at the feature-definition level, allowing different groups to compute mathematically comparable values from the same image [4, 5]. Yet a well-known practical obstacle remains: the features produced by a typical pipeline are heavily collinear. Researchers routinely deal with this by pruning highly correlated pairs, by projecting onto principal components, or by using regularised model fits — operations that are performed afresh for every new dataset, and that therefore yield slightly different feature subsets every time.

Two assumptions implicit in this routine pruning deserve scrutiny. The first is that redundancy is a property of the study cohort. The second is that the specific feature retained at the end of pruning is essentially interchangeable with the ones discarded. Both assumptions become questionable as soon as one notices that highly correlated pairs tend to involve features with overlapping mathematical definitions: a long-run emphasis from the gray-level run-length matrix is almost identical to a large-dependence emphasis from the gray-level dependence matrix; a first-order variance is closely related to a sum-of-squares descriptor from the gray-level co-occurrence matrix; voxel volume, mesh volume, and surface area encode highly correlated views of the same morphology. If these correlations arise because pairs of features quantify the same underlying image property, they should not be dataset-specific. They should be universal.

In this work we test that hypothesis directly. We re-analyse the recently published Radiomics Atlas Dataset of normal Abdominal and Pelvic CT (RADAPT), which provides a uniformly extracted radiomics description of 53 anatomical structures in 531 healthy young adults using a standardised PyRadiomics pipeline [6]. Because RADAPT spans bones, muscles, vessels, and parenchymal organs in the same subjects, it is well suited to ask whether feature–feature correlations survive simultaneously across all of these tissue categories. We restrict the analysis to the original (non-filtered) PyRadiomics features — the descriptors computed directly on the unmodified image — to keep the discussion grounded in features that have clear single-image meaning, and we use a strict, global-intersection criterion: a feature pair is reported as universally correlated only if its Spearman |ρ| exceeds a fixed threshold in every structure (each correlation estimated across the full patient sample).

The pipeline addresses several recurring concerns in the field. First, the universally correlated communities give an interpretable, anatomy-independent basis for dimensionality reduction: choosing one representative per community is mathematically justified rather than ad hoc. Second, mapping which features are interchangeable helps reconcile the well-known phenomenon in which two radiomics studies of the same disease pick almost entirely different feature subsets and yet reach overlapping conclusions — the subsets often come from the same universal communities, and so encode the same image information. Third, restricting predictive models to within-community representatives is a concrete step toward organ-agnostic radiomics, because by construction the retained features behave consistently in muscles, vessels, parenchyma, and bone. Where it aids interpretation, we additionally relate each community to the radiological vocabulary that recent work has begun to attach to individual radiomics features — heterogeneity, signal level (hyper-/hypo-intensity), texture coarseness, and lesion size/compactness — to make explicit what clinical property each community summarises [7].

## 2. Materials and Methods

### 2.1. Dataset

All analyses used the publicly available RADAPT dataset (https://github.com/eliskape/Radiomics-Atlas), released under a Creative Commons CC-BY-NC-ND licence [6]. RADAPT contains radiomics features extracted from abdominal and pelvic CT scans of 531 young adults (281 male, 250 female; mean age 26.8 ± 5.2 years) without previously known disease, who underwent CT scanning for emergency indications. Image acquisition used 64-slice GE or Siemens scanners with 120 kV tube voltage, 0.4-s rotation time, a 500 × 500 mm field of view, and 3.75-mm reconstructed slice thickness. Anatomical structures were segmented automatically with the TotalSegmentator deep-learning model deployed in 3D Slicer 5.2.1, and abnormal structures, identifiable lesions, and structures with variable filling (gastrointestinal tract, urinary bladder) were excluded by two radiologists [6, 8]. Radiomics features were extracted with PyRadiomics using a uniform bin width of 25 HU and voxel resampling to 4 × 4 × 4 mm^3^ [5].

We used only the non-contrast-enhanced series (n = 526 of the 531-subject atlas) and only the original (non-filtered) feature classes; wavelet and Laplacian-of-Gaussian features were excluded. Restricting to non-contrast images removes the confound of contrast timing on intensity-derived features, and restricting to original features keeps every value interpretable as a single-image statistic without an additional image transform. We organised the 53 anatomical structures into four broad anatomical categories (Table 1).

**Table 1.** Anatomical grouping of the 53 RADAPT structures used in this study.

| Anatomical group | Representative organs / structures included |
| --- | --- |
| Bones | Lower thoracic and lumbar vertebrae, sacrum, lower ribs, pelvic bones (ilium, ischium, pubis), proximal femurs |
| Muscles | Iliopsoas, gluteus maximus / medius / minimus, autochthonous (paraspinal) musculature |
| Vessels | Aorta, inferior vena cava, common iliac arteries and veins, portal vein |
| Parenchymal organs | Liver, spleen, pancreas, both kidneys, both adrenal glands, gallbladder |

### 2.2. Pre-processing

Each anatomical structure in RADAPT is distributed as one Excel file per structure; rows are patients and columns are PyRadiomics features. We loaded every such file independently, dropped the patient-identifier column, and retained only columns whose name began with “original_”. To make features computed on different organs and patients directly comparable while preserving the within-file ranking, every feature was z-score-standardised within its file (mean 0, standard deviation 1, with zero standard deviations replaced by 1 to avoid division by zero). We deliberately did not pool data across files before standardisation, so that no organ-specific scale could artificially inflate or shrink correlations. Because correlations were computed on ranks (Spearman), this standardisation does not itself affect the correlation values; it is retained for transparency and to keep the per-file feature matrices on a common scale for visualisation.

### 2.3. Per-file correlation graphs

For each structure file we computed the absolute Spearman rank correlation matrix across patients on the standardised features. Spearman correlation was preferred over Pearson because it is invariant to monotone transforms of the features and is therefore robust to the heavy-tailed distributions that are common in radiomics descriptors; as a consequence, feature pairs related by any monotone transform (for example standard deviation and variance) attain |ρ| = 1 by construction. Feature pairs with |ρ| greater than or equal to a fixed threshold τ were retained as edges. We swept τ ∈ {0.70, 0.80, 0.90} to characterise how the community structure tightens as the criterion becomes more stringent.

### 2.4. Universal correlation graph and community detection

The central methodological step is the construction of a universal correlation graph by intersection. Let El be the set of edges retained in structure file k. The universal edge set at threshold τ is

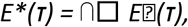

so that a feature pair appears in E*(τ) only if it is jointly highly correlated in every anatomical structure examined (each correlation estimated across the full patient sample). This is a far stricter condition than averaging correlations across the dataset and then thresholding, because a single structure in which the pair is uncorrelated removes the edge permanently; the four anatomical categories therefore enter the analysis as the breadth of tissue over which an edge must survive, not as a separate averaging step. We built the universal graph in NetworkX [9] and enumerated stable feature communities as its maximal cliques of size ≥ 2 using the Bron–Kerbosch algorithm [10]. Reporting cliques rather than connected components preserves the requirement that every pair within a community is mutually highly correlated, not merely connected by some chain of pairwise correlations.

### 2.5. Reproducibility analyses

We assessed reproducibility in two complementary ways. First, we re-implemented the complete pipeline independently in R (using the readxl, writexl, and igraph [11] packages) and verified that, for every threshold, the set of stable communities was identical to that produced by the Python implementation; this checks implementation correctness and freedom from library- or floating-point–ordering artefacts, not external validity. Second, we performed a random-half stability test in R: with a fixed seed (123), each structure file was split row-wise (i.e. by patient) into two non-overlapping halves; the full intersection-and-cliques pipeline was re-run on each half-cohort independently; and the resulting communities were compared back to those obtained on the full dataset. This procedure was repeated for five independent random partitions, yielding ten sub-cohort runs per threshold. For each run we counted the number of stable communities recovered exactly (i.e. with the same feature membership as the corresponding full-data community).

### 2.6. Software and reproducibility

The Python pipeline used pandas 2.x, NumPy, scikit-learn (only for StandardScaler), NetworkX, and seaborn; the R pipeline used readxl, writexl, and igraph. Network visualisations were produced in R using the ggraph layer of the tidygraph framework. All code and intermediate result tables are available from the authors on request.

## 3. Results

### 3.1. Number of universally correlated communities scales smoothly with threshold

Applying the global-intersection pipeline to RADAPT yielded a strictly nested sequence of universal correlation graphs across the three thresholds. As the Spearman threshold tightened, both the number of surviving universal edges and the number of stable cliques decreased monotonically, but the structure did not collapse: even at the strictest threshold of |ρ| ≥ 0.90, fourteen distinct multi-feature communities remained that were universal across every patient and every anatomical structure (Table 2).

**Table 2.**
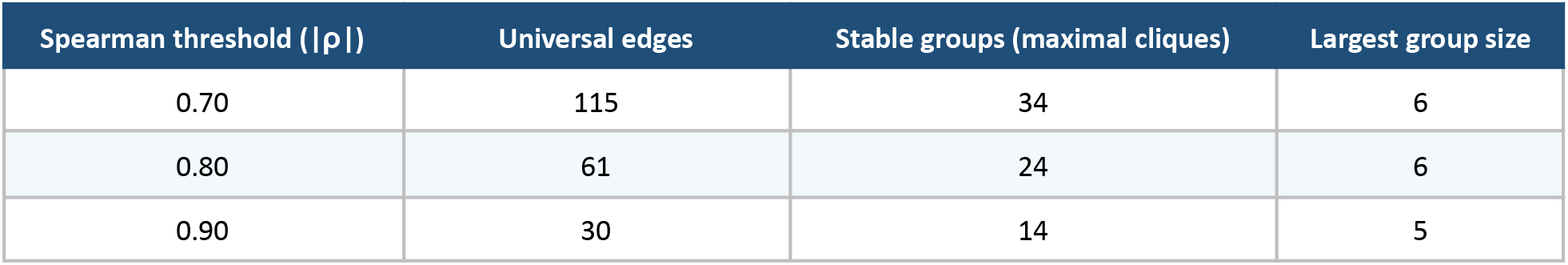
Number of universal edges and stable feature communities at each correlation threshold.

| Spearman threshold ($\vert \rho\vert $) | Universal edges | Stable groups (maximal cliques) | Largest group size |
| --- | --- | --- | --- |
| 0.70 | 115 | 34 | 6 |
| 0.80 | 61 | 24 | 6 |
| 0.90 | 30 | 14 | 5 |

The fact that fourteen communities survive the strictest test, and that the largest of them contains five mutually almost-perfectly-correlated features, is the central finding of this work. It is incompatible with the view that radiomics feature redundancy is a per-cohort artefact: edges that vary across patients, across muscles versus parenchyma, or across bones versus vessels are eliminated by the intersection, and only the truly anatomy- and patient-independent edges remain.

### 3.2. Nested structure of the communities across thresholds

Inspecting the three universal graphs together reveals a clean nesting. Every clique at τ = 0.90 is contained in a clique at τ = 0.80, and every clique at τ = 0.80 is contained in a clique at τ = 0.70 (Figure 1). Some communities — for example the long-run / large-dependence cluster of GLRLM and GLDM descriptors, and the volume / surface shape cluster — remain almost intact across all three thresholds, indicating that the features in them are essentially numerical aliases of one another. Others, such as the variance / sum-of-squares family, grow at lower thresholds by absorbing additional GLCM-derived descriptors (for example ClusterProminence, ClusterTendency, and DifferenceVariance), reflecting a more graded form of redundancy.

**Figure 1.**
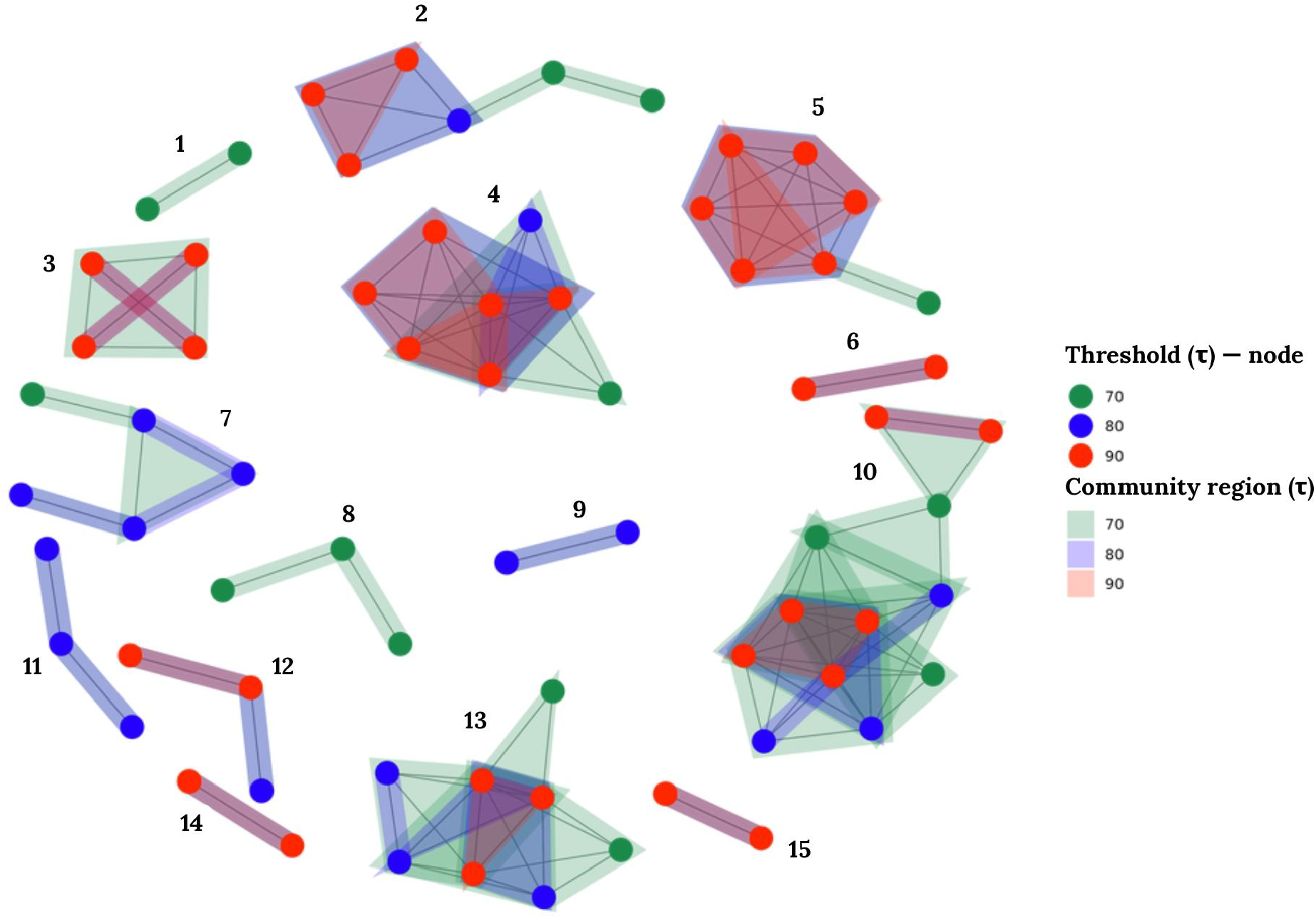
Universal radiomics correlation graph at three Spearman thresholds, overlaid on a single circular layout. Nodes are PyRadiomics original features that participate in at least one universal community; edges connect features whose absolute Spearman correlation exceeds the threshold in every patient and every anatomical structure in RADAPT. Green = τ ≥ 0.70; blue = τ ≥ 0.80; red = τ ≥ 0.90. Cliques shrink monotonically as the threshold tightens but do not disappear, showing that a core of universal feature redundancy is robust to even a very strict correlation criterion.

Figure 2 shows a detailed view of community #13 from Figure 1 as a representative example, and illustrates how its clique structure changes as the correlation threshold is varied.

**Figure 2.**
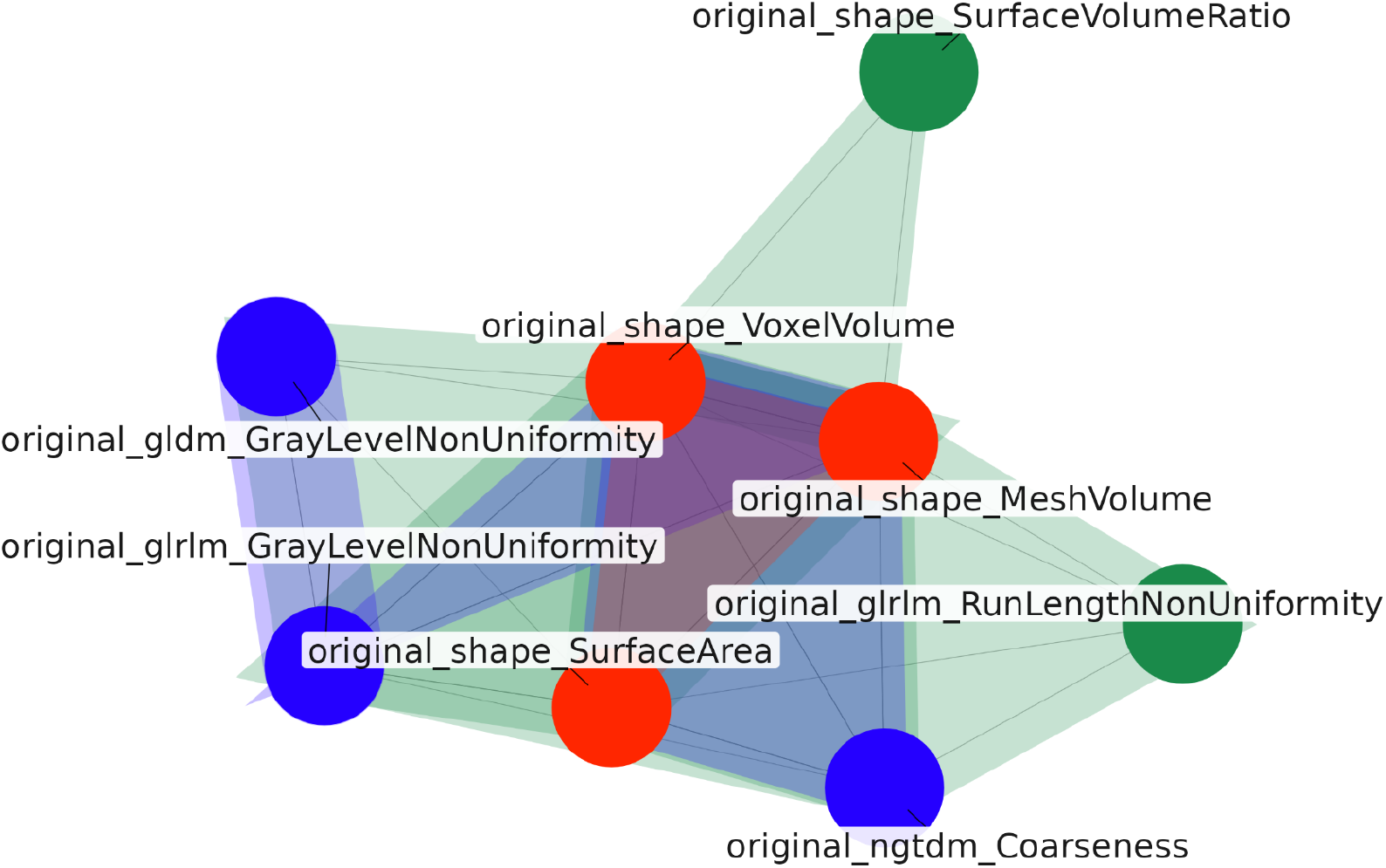
Detailed view of community #13 from Figure 1. The community spans four PyRadiomics feature classes — shape (VoxelVolume, MeshVolume, SurfaceArea, SurfaceVolumeRatio), GLDM (GrayLevelNonUniformity), GLRLM (GrayLevelNonUniformity, RunLengthNonUniformity), and NGTDM (Coarseness) — with a core subset forming a clique already at τ = 0.90 (red) and additional features joining as the threshold is relaxed to τ = 0.80 (blue) and τ = 0.70 (green). Colors and edge conventions are as in Figure 1.

### 3.3. Interpretation of the largest universal communities

Although our analysis is purely statistical, the communities map cleanly onto well-known mathematical relationships between PyRadiomics descriptors, and each maps in turn onto a single radiological property — the vocabulary radiologists already use to describe a region of interest [7]. Six broad themes emerge consistently across the three thresholds.

- Long-run / large-dependence (coarse) texture. GLRLM LongRunEmphasis, GLRLM RunPercentage, GLRLM RunVariance, GLRLM ShortRunEmphasis (inversely), GLRLM RunLengthNonUniformityNormalized, and GLDM LargeDependenceEmphasis all summarise the prevalence of long, spatially extended runs of similar intensity, i.e. coarse, homogeneous texture. They cluster together at every threshold. Radiologically this is the coarseness/homogeneity axis used to separate uniform tissue from finely heterogeneous lesions.
- High gray-level texture. GLCM Autocorrelation, GLCM JointAverage, GLCM SumAverage, GLDM HighGrayLevelEmphasis, GLRLM HighGrayLevelRunEmphasis, and GLRLM ShortRunHighGrayLevelEmphasis all increase with the mean image intensity within the ROI and form a single tight community. This is the quantitative substrate of overall signal level — the hyper-versus hypo-intensity reading that drives suspicion on, for example, high-b-value DWI.
- Variance-like intensity dispersion. First-order Variance, GLCM ClusterTendency, GLCM SumSquares, GLDM GrayLevelVariance, and GLRLM GrayLevelVariance are all variance estimators on the discretised intensity histogram, and they remain a single clique even at |ρ| ≥ 0.90. At lower thresholds the community widens to include GLCM ClusterProminence, GLCM DifferenceVariance, and first-order MeanAbsoluteDeviation. Intensity dispersion is the direct quantitative reading of lesion heterogeneity, the property that most consistently raises radiological suspicion.
- Volume and surface shape. Mesh volume, voxel volume, and surface area form a stable triangle at every threshold; at τ = 0.70 the community grows by absorbing NGTDM Coarseness and several GLRLM / GLDM non-uniformity descriptors, all of which scale with absolute ROI size. Size is itself a clinical axis — for instance, the 1.5-cm threshold that separates higher from lower PI-RADS categories — and SurfaceVolumeRatio adds a compactness/shape reading on the same community.
- Low gray-level texture. GLDM LowGrayLevelEmphasis, GLRLM LowGrayLevelRunEmphasis, GLRLM ShortRunLowGrayLevelEmphasis, and GLSZM LowGrayLevelZoneEmphasis cluster together, mirroring the high gray-level community on the opposite end of the intensity range — the quantitative substrate of hypo-intensity.
- Local homogeneity and energy / entropy duals. GLCM Id ↔ GLCM Idm, GLCM JointEnergy ↔ GLCM MaximumProbability, first-order Entropy ↔ first-order Uniformity, and first-order Energy ↔ first-order TotalEnergy all appear as universal two-feature cliques. These differ in kind from the multi-family communities above: they are (near-)algebraic identities. TotalEnergy is Energy multiplied by the voxel volume, so under the fixed 4 × 4 × 4 mm^3^ resampling used in RADAPT the two are perfectly correlated by construction; Entropy and Uniformity are competing summaries of the same histogram and are near-deterministically related; Id and Idm differ only in a distance weighting. Their universality is therefore expected and carries no biological content.

This last point organises the whole result. The two-feature duals are universal trivially, because the IBSI definitions encode them as the same quantity in different notation; the scientifically informative finding is the set of cross-matrix-family communities — variance estimators drawn from first-order, GLCM, GLDM, and GLRLM; the high– and low–gray-level families spanning three matrices; and the long-run / large-dependence family spanning GLRLM and GLDM — which are not exact identities yet remain locked together in every structure and every patient subset. Together these themes account for the majority of the cliques in Tables 3–5: rather than thirty-four independent forms of redundancy at τ = 0.70, RADAPT exhibits a handful of underlying image properties that the IBSI feature definitions express in slightly different ways.

**Table 3.** Stable feature communities at Spearman |ρ| ≥ 0.70 (34 maximal cliques).

| Group | Size | Features (PyRadiomics names, 'original_' prefix omitted) |
| --- | --- | --- |
| 1 | 6 | gldm_LargeDependenceEmphasis, glrlm_LongRunEmphasis, glrlm_RunLengthNonUniformityNormalized, glrlm_RunPercentage, glrlm_RunVariance, glrlm_ShortRunEmphasis |
| 2 | 6 | firstorder_Variance, glcm_ClusterTendency, glcm_DifferenceVariance, glcm_SumSquares, gldm_GrayLevelVariance, glrlm_GrayLevelVariance |
| 3 | 6 | firstorder_Variance, glcm_ClusterProminence, glcm_ClusterTendency, glcm_SumSquares, gldm_GrayLevelVariance, glrlm_GrayLevelVariance |
| 4 | 6 | glcm_Autocorrelation, glcm_JointAverage, glcm_SumAverage, gldm_HighGrayLevelEmphasis, glrlm_HighGrayLevelRunEmphasis, glrlm_ShortRunHighGrayLevelEmphasis |
| 5 | 5 | gldm_DependenceNonUniformityNormalized, gldm_LargeDependenceEmphasis, glrlm_LongRunEmphasis, glrlm_RunPercentage, glrlm_RunVariance |
| 6 | 5 | glrlm_RunLengthNonUniformity, ngtdm_Coarseness, shape_MeshVolume, shape_SurfaceArea, shape_VoxelVolume |
| 7 | 5 | glrlm_GrayLevelNonUniformity, ngtdm_Coarseness, shape_MeshVolume, shape_SurfaceArea, shape_VoxelVolume |
| 8 | 5 | gldm_GrayLevelNonUniformity, glrlm_GrayLevelNonUniformity, shape_MeshVolume, shape_SurfaceArea, shape_VoxelVolume |
| 9 | 5 | firstorder_MeanAbsoluteDeviation, firstorder_Variance, glcm_ClusterTendency, glcm_SumSquares, gldm_GrayLevelVariance |
| 10 | 5 | firstorder_Variance, glcm_Contrast, glcm_SumSquares, gldm_GrayLevelVariance, glrlm_GrayLevelVariance |
| 11 | 5 | firstorder_MeanAbsoluteDeviation, firstorder_Variance, glcm_Contrast, glcm_SumSquares, gldm_GrayLevelVariance |
| 12 | 4 | gldm_DependenceVariance, gldm_LargeDependenceEmphasis, glrlm_LongRunEmphasis, glrlm_RunVariance |
| 13 | 4 | gldm_LowGrayLevelEmphasis, glrlm_LowGrayLevelRunEmphasis, glrlm_ShortRunLowGrayLevelEmphasis, glszm_LowGrayLevelZoneEmphasis |
| 14 | 4 | gldm_SmallDependenceEmphasis, glszm_LargeAreaEmphasis, glszm_ZonePercentage, glszm_ZoneVariance |
| 15 | 3 | glcm_DifferenceAverage, glcm_Id, glcm_Idm |
| 16 | 3 | shape_MeshVolume, shape_SurfaceVolumeRatio, shape_VoxelVolume |
| 17 | 3 | glcm_JointEnergy, glcm_JointEntropy, glcm_SumEntropy |
| 18 | 3 | firstorder_MeanAbsoluteDeviation, glcm_Contrast, glcm_DifferenceAverage |
| 19 | 2 | gldm_SmallDependenceLowGrayLevelEmphasis, glszm_SmallAreaLowGrayLevelEmphasis |
| 20 | 2 | glszm_LowGrayLevelZoneEmphasis, glszm_SmallAreaLowGrayLevelEmphasis |
| 21 | 2 | glcm_JointEnergy, glcm_MaximumProbability |
| 22 | 2 | glszm_HighGrayLevelZoneEmphasis, glszm_SmallAreaHighGrayLevelEmphasis |
| 23 | 2 | glcm_DifferenceEntropy, glcm_SumEntropy |
| 24 | 2 | firstorder_Energy, firstorder_TotalEnergy |
| 25 | 2 | firstorder_InterquartileRange, firstorder_RobustMeanAbsoluteDeviation |
| 26 | 2 | glszm_SizeZoneNonUniformityNormalized, glszm_SmallAreaEmphasis |
| 27 | 2 | gldm_SmallDependenceHighGrayLevelEmphasis, glrlm_ShortRunHighGrayLevelEmphasis |
| 28 | 2 | firstorder_Entropy, glrlm_GrayLevelNonUniformityNormalized |
| 29 | 2 | gldm_LargeDependenceLowGrayLevelEmphasis, glszm_LargeAreaLowGrayLevelEmphasis |
| 30 | 2 | glszm_GrayLevelVariance, ngtdm_Complexity |
| 31 | 2 | firstorder_Range, ngtdm_Complexity |
| 32 | 2 | firstorder_Entropy, firstorder_Uniformity |
| 33 | 2 | firstorder_Mean, firstorder_Median |
| 34 | 2 | firstorder_90Percentile, firstorder_Mean |

### 3.4. Full enumeration of stable communities

Tables 3, 4, and 5 list every stable feature community at the three thresholds. To keep the tables compact we omit the “original_” prefix from feature names.

**Table 4.** Stable feature communities at Spearman |ρ| ≥ 0.80 (24 maximal cliques).

| Group | Size | Features (PyRadiomics names, 'original_' prefix omitted) |
| --- | --- | --- |
| 1 | 6 | glcm_Autocorrelation, glcm_JointAverage, glcm_SumAverage, gldm_HighGrayLevelEmphasis, glrlm_HighGrayLevelRunEmphasis, glrlm_ShortRunHighGrayLevelEmphasis |
| 2 | 6 | gldm_LargeDependenceEmphasis, glrlm_LongRunEmphasis, glrlm_RunLengthNonUniformityNormalized, glrlm_RunPercentage, glrlm_RunVariance, glrlm_ShortRunEmphasis |
| 3 | 5 | firstorder_Variance, glcm_ClusterTendency, glcm_SumSquares, gldm_GrayLevelVariance, glrlm_GrayLevelVariance |
| 4 | 4 | ngtdm_Coarseness, shape_MeshVolume, shape_SurfaceArea, shape_VoxelVolume |
| 5 | 4 | gldm_DependenceNonUniformityNormalized, gldm_LargeDependenceEmphasis, glrlm_LongRunEmphasis, glrlm_RunVariance |
| 6 | 4 | gldm_LowGrayLevelEmphasis, glrlm_LowGrayLevelRunEmphasis, glrlm_ShortRunLowGrayLevelEmphasis, glszm_LowGrayLevelZoneEmphasis |
| 7 | 3 | glrlm_GrayLevelNonUniformity, shape_MeshVolume, shape_VoxelVolume |
| 8 | 2 | glcm_Id, glcm_Idm |
| 9 | 2 | glcm_JointEnergy, glcm_MaximumProbability |
| 10 | 2 | gldm_SmallDependenceEmphasis, glszm_ZonePercentage |
| 11 | 2 | firstorder_Entropy, firstorder_Uniformity |
| 12 | 2 | glcm_DifferenceVariance, glcm_SumSquares |
| 13 | 2 | glcm_JointEntropy, glcm_SumEntropy |
| 14 | 2 | glszm_HighGrayLevelZoneEmphasis, glszm_SmallAreaHighGrayLevelEmphasis |
| 15 | 2 | firstorder_InterquartileRange, firstorder_RobustMeanAbsoluteDeviation |
| 16 | 2 | firstorder_Energy, firstorder_TotalEnergy |
| 17 | 2 | glcm_JointEnergy, glcm_JointEntropy |
| 18 | 2 | glszm_SizeZoneNonUniformityNormalized, glszm_SmallAreaEmphasis |
| 19 | 2 | firstorder_Entropy, glrlm_GrayLevelNonUniformityNormalized |
| 20 | 2 | firstorder_Mean, firstorder_Median |
| 21 | 2 | firstorder_90Percentile, firstorder_Mean |
| 22 | 2 | gldm_GrayLevelNonUniformity, glrlm_GrayLevelNonUniformity |
| 23 | 2 | firstorder_MeanAbsoluteDeviation, glcm_SumSquares |
| 24 | 2 | glszm_LargeAreaEmphasis, glszm_ZoneVariance |

**Table 5.** Stable feature communities at Spearman |ρ| ≥ 0.90 (14 maximal cliques).

| Group | Size | Features (PyRadiomics names, 'original_' prefix omitted) |
| --- | --- | --- |
| 1 | 5 | glcm_Autocorrelation, glcm_JointAverage, glcm_SumAverage, gldm_HighGrayLevelEmphasis, glrlm_HighGrayLevelRunEmphasis |
| 2 | 5 | gldm_LargeDependenceEmphasis, glrlm_LongRunEmphasis, glrlm_RunLengthNonUniformityNormalized, glrlm_RunPercentage, glrlm_ShortRunEmphasis |
| 3 | 4 | firstorder_Variance, glcm_SumSquares, gldm_GrayLevelVariance, glrlm_GrayLevelVariance |
| 4 | 4 | gldm_LargeDependenceEmphasis, glrlm_LongRunEmphasis, glrlm_RunPercentage, glrlm_RunVariance |
| 5 | 3 | shape_MeshVolume, shape_SurfaceArea, shape_VoxelVolume |
| 6 | 3 | gldm_LowGrayLevelEmphasis, glrlm_LowGrayLevelRunEmphasis, glrlm_ShortRunLowGrayLevelEmphasis |
| 7 | 3 | gldm_HighGrayLevelEmphasis, glrlm_HighGrayLevelRunEmphasis, glrlm_ShortRunHighGrayLevelEmphasis |
| 8 | 2 | glcm_Id, glcm_Idm |
| 9 | 2 | gldm_SmallDependenceEmphasis, glszm_ZonePercentage |
| 10 | 2 | firstorder_Entropy, firstorder_Uniformity |
| 11 | 2 | firstorder_InterquartileRange, firstorder_RobustMeanAbsoluteDeviation |
| 12 | 2 | firstorder_Energy, firstorder_TotalEnergy |
| 13 | 2 | glszm_SizeZoneNonUniformityNormalized, glszm_SmallAreaEmphasis |
| 14 | 2 | glszm_LargeAreaEmphasis, glszm_ZoneVariance |

### 3.5. The communities are reproducible under resampling and cross-language re-implementation

Two independent reproducibility checks support the universality interpretation. First, an R re-implementation of the entire pipeline (using the igraph package for graph construction and clique enumeration) returned exactly the same set of communities as the Python implementation at all three thresholds, confirming that the result is not an artefact of any particular library or floating-point ordering.

Second, in the random-half stability experiment, each input file was randomly partitioned by patient into two non-overlapping halves and the complete intersection-plus-cliques pipeline was re-run on each half. Across five independent partitions (ten sub-cohort runs per threshold), the number of stable communities recovered in a random half was always close to the full-data value, and a large majority of those communities matched the full-data communities exactly in their feature membership (Table 6).

**Table 6.** Recovery of universal feature communities under random-half resampling (5 partitions × 2 halves = 10 sub-cohort runs per threshold). Mean ± SD across runs.

| Threshold | Groups (full data) | Groups in random half (mean ± SD) | Exactly-recovered groups (mean ± SD) | Recovery rate |
| --- | --- | --- | --- | --- |
| 0.70 | 34 | 33.6 ± 2.2 | 27.7 ± 1.8 | 81.5 % |
| 0.80 | 24 | 24.0 ± 1.7 | 20.8 ± 1.4 | 86.7 % |
| 0.90 | 14 | 15.3 ± 1.1 | 11.3 ± 1.0 | 80.7 % |

The departures from exact recovery have a simple and consistent explanation. In a half-sample the correlation of each feature pair is estimated from roughly half as many patients, so its sampling error is larger and pairs whose true |ρ| lies just above or below τ can flip across the threshold. Because communities are defined as maximal cliques, a single flipped edge does not perturb the membership at random: dropping one within-clique edge splits a clique into two smaller cliques, which can make the half-data community count exceed the full-data count (as seen at τ = 0.90, where 15.3 ± 1.1 groups were found against 14 in the full data), while gaining one edge can let a clique absorb an adjacent feature.

Consistent with this, the non-exact matches were almost always a single missing or extra feature — typically one sitting on the boundary between two adjacent cliques — rather than a structurally different grouping. The pattern indicates that the universal communities reflect a stable underlying redundancy structure that is already well sampled by roughly half of the patients, and that the small run-to-run differences are finite-sample threshold effects rather than evidence of instability.

## 4. Discussion

We have shown that a substantial portion of the feature–feature redundancy that characterises radiomics is not a property of any one cohort, organ, or disease, but a property of the feature extractor itself as it operates on real medical images. Even under a strict global-intersection criterion that requires high pairwise correlation in every anatomical structure and (through resampling) every patient subset simultaneously, fourteen distinct multi-feature communities survive at |ρ| ≥ 0.90 across the 526 non-contrast examinations and four anatomical categories of the RADAPT atlas. The communities are recovered with > 80 % exact-match rates under random half-sample resampling, and an independent R re-implementation reproduces them identically. These are the properties one expects of features that share an underlying definitional or image-statistical meaning, not of features that happen to be correlated in one cohort.

The community structure is also interpretable, and it is important to separate two layers within it. The two-feature cliques — GLCM Id ↔ Idm, GLCM JointEnergy ↔ MaximumProbability, first-order Entropy ↔ Uniformity, first-order Energy ↔ TotalEnergy — are near-algebraic identities: Energy and TotalEnergy differ only by the (here constant) voxel volume, and the others are competing summaries of a single histogram. Their universality is therefore guaranteed by the IBSI definitions and is not, in itself, a discovery; it does, however, serve as a positive control that the pipeline recovers known identities. The non-trivial result lies in the larger cross-matrix-family communities: variance-like estimators of the intensity histogram drawn from first-order, GLCM, GLDM, and GLRLM (first-order Variance, GLCM SumSquares, GLCM ClusterTendency, GLDM and GLRLM GrayLevelVariance); summaries of mean intensity spanning three matrices (GLCM JointAverage, GLCM SumAverage, GLCM Autocorrelation, GLDM and GLRLM HighGrayLevelEmphasis); descriptors of long, homogeneous runs of similar intensity (the joint GLRLM long-run / GLDM large-dependence family); and volumetric shape (mesh volume, voxel volume, surface area). These are not exact identities, yet they behave as identities across very different anatomy — that is the empirical content of the paper. Each of these communities also corresponds to a single radiological reading — heterogeneity, signal level, coarseness, and lesion size respectively — which is the vocabulary recent feature-dictionary work has begun to attach to individual descriptors [7], so that the statistical communities can be named in clinically meaningful terms.

The most direct practical consequence is that the original PyRadiomics feature space can be compressed by roughly an order of magnitude with negligible loss of distinct information. Selecting a single representative — for example, the most interpretable or lowest-variance member of each community — yields a curated set of around fourteen features at τ = 0.90, or around twenty-four at τ = 0.80, that together span every dimension on which RADAPT features differ universally. Because the communities were defined without reference to any disease label, this reduction is unsupervised and does not bias subsequent model fitting; it simply removes mathematical aliases that would otherwise inflate the effective number of tests, destabilise penalised regressions, and degrade interpretability.

A second, less direct consequence relates to the interpretation of existing literature. It is well known that two radiomics studies addressing the same clinical question often select almost disjoint subsets of features and yet report similar predictive performance and similar biological narratives [3, 12]. Our results suggest that this is largely expected behaviour: in many cases the disjoint feature subsets are different draws from the same universal communities, so the studies are effectively measuring the same six or seven underlying themes and merely labelling them with different IBSI names. Reporting which universal community each selected feature belongs to — rather than only the feature name — would make such studies directly comparable and would clarify when a discrepancy is genuine.

Third, restricting predictive models to within-community representatives is a concrete step toward what has been called organ-agnostic radiomics. By construction, the universal communities are stable across bones, muscles, vessels, and parenchymal organs, so a model built only on community representatives is forced to rely on signal that does not depend on the background organ phenotype. This is the opposite of what happens when an unconstrained model trained on a single organ exploits organ-specific feature behaviour and then fails to generalise when deployed elsewhere [13, 14].

### Limitations

Several limitations should be acknowledged. First, RADAPT consists exclusively of CT images from young adults aged 17–36 years scanned for emergency indications and reported as normal, with two specific scanner vendors, a uniform reconstruction slice thickness of 3.75 mm, and PyRadiomics extraction at a 25-HU bin width and 4 × 4 × 4 mm^3^ voxel resampling [6]. Because feature values — and in particular volume-confounded features such as Energy and TotalEnergy — depend on these acquisition and extraction settings, the precise set of universal communities reported here is, strictly speaking, conditional on these settings. We expect the qualitative themes (variance, long runs, high / low gray level, volume / surface, homogeneity / entropy duals) to be robust to such changes, because they reflect relationships between feature definitions; but the precise membership of individual cliques should be re-checked when extending the analysis to other modalities, age groups, scanners, or pathological cohorts. In particular, because RADAPT contains only normal structures, communities that depend on the presence of focal lesions cannot be observed here, and the transfer of these communities to diseased tissue is an explicit hypothesis for future work rather than an established result.

Second, we restricted attention to the original (non-filtered) PyRadiomics feature classes. Wavelet and Laplacian-of-Gaussian transforms generate hundreds of additional features whose redundancy structure is likely to be even richer and is an obvious target for future work. Third, we used the maximal-clique definition of a community, which guarantees mutual pairwise correlation but can split larger groups when a single edge falls just below threshold; alternative community-detection algorithms (e.g. Louvain or Leiden on the full universal graph) may produce coarser groupings that are easier to use as drop-in replacements for the original feature set. Fourth, the random-half resampling test demonstrates stability of the communities at the level of the RADAPT cohort, but external validation on a second public atlas would be required to certify that the communities transfer between datasets.

## 5. Conclusion

A substantial part of the feature redundancy that researchers routinely encounter in radiomics is universal: it persists across patients and across very different anatomical structures because it reflects shared mathematical and image-statistical realities of how radiomics features summarise an image. Using a strict global-intersection criterion on the RADAPT CT atlas, we identified 34, 24, and 14 stable, mutually correlated feature communities at Spearman thresholds of 0.70, 0.80, and 0.90 respectively, distinguishing trivial near-algebraic duals from the non-trivial cross-matrix-family communities that constitute the real finding. The communities are reproducible across random sub-cohorts and across independent Python and R implementations, and they correspond to a small number of clearly interpretable themes that can be named in radiological terms. We argue that these universal communities provide a principled basis for unsupervised dimensionality reduction, for retrospectively reconciling apparently different feature selections across radiomics studies, and for building more generalisable, organ-agnostic radiomics models.

## Data Availability

All data produced are available online at: https://github.com/eliskape/Radiomics-Atlas

## Abbreviations

CT,: computed tomography;
GLCM,: gray-level co-occurrence matrix;
GLDM,: gray-level dependence matrix;
GLRLM,: gray-level run-length matrix;
GLSZM,: gray-level size-zone matrix;
NGTDM,: neighbouring gray-tone difference matrix;
IBSI,: Image Biomarker Standardisation Initiative;
ROI,: region of interest;
RADAPT,: Radiomics Atlas Dataset of normal Abdominal and Pelvic CT.

## Data and code availability

The RADAPT dataset is publicly available at https://github.com/eliskape/Radiomics-Atlas under a CC-BY-NC-ND licence [6]. Code, derived universal correlation graphs, and the full per-threshold community tables produced in this study are available from the authors on reasonable request.

## Declarations

Conflict of interest. The authors declare that they have no competing interests.

Ethics approval. This study used a publicly available, fully anonymised radiomics dataset (RADAPT) released under CC-BY-NC-ND. No new patient data were collected. No additional ethics approval was therefore required.

## References

[1] P. Lambin, E. Rios-Velazquez and R. Leijenaar, “Radiomics: extracting more information from medical images using advanced feature analysis,” Eur J Cancer, vol. 48, no. 4, pp. 441–446, 2012.

[2] R. Gillies, P. Kinahan and H. Hricak, “Radiomics: images are more than pictures, they are data,” Radiology, vol. 278, no. 2, pp. 563–577, 2016.

[3] H. Aerts, E. Velazquez and R. Leijenaar, “Decoding tumour phenotype by noninvasive imaging using a quantitative radiomics approach,” Nat Commun, vol. 5, p. 4006, 2014.

[4] A. Zwanenburg, M. Vallieres and M. Abdalah, “The Image Biomarker Standardization Initiative: standardized quantitative radiomics for high-throughput image-based phenotyping,” Radiology, vol. 295, no. 2, pp. 328–338, 2020.

[5] J. van Griethuysen, A. Fedorov and C. Parmar, “Computational radiomics system to decode the radiographic phenotype,” Cancer Res, vol. 77, no. 21, pp. e104–e107, 2017.

[6] E. Kapetanou, S. Malamas, D. Leventis, A. Karantanas and M. Klontzas, “Developing a Radiomics Atlas Dataset of normal Abdominal and Pelvic computed Tomography (RADAPT),” J Digit Imaging Inform Med, vol. 37, pp. 1273–1281, 2024.

[7] M. Salmanpour, S. Amiri and S. Gharibi, “Biological and radiological dictionary of radiomics features: addressing understandable AI issues in personalized prostate cancer (dictionary version PM1.0),” 2024.

[8] J. Wasserthal, H. Breit and M. Meyer, “TotalSegmentator: robust segmentation of 104 anatomical structures in CT images,” Radiol Artif Intell, vol. 5, no. 5, p. e230024, 2023.

[9] A. Hagberg, D. Schult and P. Swart, “Exploring network structure, dynamics, and function using NetworkX,” in Proceedings of the 7th Python in Science Conference (SciPy 2008), 2008.

[10] C. Bron and J. Kerbosch, “Algorithm 457: finding all cliques of an undirected graph,” Commun ACM, vol. 16, no. 9, pp. 575–577, 1973.

[11] G. Csardi and T. Nepusz, “The igraph software package for complex network research,” InterJournal Complex Syst, vol. 1695, pp. 1–9, 2006.

[12] R. Berenguer, M. Pastor-Juan and J. Canales-Vazquez, “Radiomics of CT features may be nonreproducible and redundant: influence of CT acquisition parameters,” Radiology, vol. 288, no. 2, pp. 407–415, 2018.

[13] J. Park, S. Park, H. Kim and H. Kim, “Reproducibility and generalizability in radiomics modeling: possible strategies in radiologic and statistical perspectives,” Korean J Radiol, vol. 20, no. 7, pp. 1124–1137, 2019.

[14] A. Traverso, L. Wee, A. Dekker and R. Gillies, “Repeatability and reproducibility of radiomic features: a systematic review,” Int J Radiat Oncol Biol Phys, vol. 102, no. 4, pp. 1143–1158, 2018.

